# Knowledge, attitude and practice toward COVID-19 among healthcare workers in public health facilities, Eastern Ethiopia

**DOI:** 10.1101/2021.01.02.21249137

**Authors:** Alinoor Mohamed Farah, Tahir Yousuf Nour, Muse Obsiye, Mowlid Aqil Adan, Omar Moeline Ali, Muktar Arab Hussein, Abdullahi Bedel Budul, Muktar Omer, Fentabil Getnet

## Abstract

On 13 March 2020, Ethiopia reported the first confirmed case of COVID-19 in Addis Ababa. COVID-19 is likely to overwhelm an already fragile health-care delivery system and reduce the availability of services for endemic health concerns such as malaria and diarrheal diseases.

Cross sectional study was conducted on heath care workers in three public health facilities in Somali region to assess knowledge, attitude and practice towards COVID-19. T-test and ANOVA were used to analyze the relationship between the dependent, and independent variables. Spearman’s correlation was used to assess the relationship between mean knowledge and attitude scores.

A vast majority of the participants were male (n = 293, 67.5%), with a mean age of 27.6 (SD: 5.3) years. The mean knowledge score was 13.7 (SD: 2.6) and the mean attitude score 10.5 (SD: 4.1). Only 45.2 % (n = 196) of the participants had a good attitude toward COVID-19. There was a negative correlation between knowledge scores, attitude scores (r=-0.295, P<0.001) and practice (r=-0.298, P<0.001).

The overall level of knowledge was good. However, the attitude and practice were relatively low. We recommend strategies for enhancing the capacity of healthcare workers to develop positive attitude and practice.

## Background

A cluster of pneumonia cases in Wuhan, China, was reported to the World Health Organization (WHO) on 31 December 2019. The cause of the pneumonia cases was identified as the 2019 novel coronavirus. On 30 January 2020, in response to this serious situation, the World Health Organization (WHO) declared it a public health emergency of international concern on January 30 and called for collaborative efforts of all countries to prevent the rapid spread of COVID-19 (Organization, 2005) and on 11 March 2020, declared COVID-19 a pandemic(Organization, 2020d). In a period of just 11 weeks from January to mid-March 2020, COVID-19 has spread rapidly from its epicenter in Wuhan City to the rest of the world. At the time of writing of current work, there are more than 25 million confirmed covid-19 cases globally and almost 950,000 associated deaths(Organization, 2020b).

COVID-19 is highly infectious, and its main clinical symptoms include fever, dry cough, fatigue, myalgia, and dyspnea. The most commonly reported symptoms are fever, cough, myalgia or fatigue, pneumonia, and complicated dyspnea, whereas less common reported symptoms include headache, diarrhea, hemoptysis, runny nose, and phlegm-producing cough(Chen et al., 2020; Control & Prevention, 2019; Huang et al., 2020; Novel, 2020). Fatality rate was higher among middle-aged and elderly patients with pre-existing medical condition (tumor surgery, cirrhosis, hypertension, coronary heart disease, diabetes, and Parkinson’s disease). The virus is also affecting the health of younger adults(Li et al., 2020).

The pandemic is adding to the burden of endemic infectious diseases that prevail in many countries with an ongoing humanitarian response, such as cholera, measles, malaria, HIV and tuberculosis. Pre-existing poor hygiene practices, poor coverage in water and sanitation services and overcrowded living conditions also augment the incidence and spread of contamination by the virus.

Ethiopia reported the first confirmed case of COVID-19 in Addis Ababa on 15 March 2020. As of 3rd of August, there were a total of 18,706 COVID-19confirmed cases and 310 associated deaths. In Somali region, the first case was identified on 24 of April 2020 and as of 3rd of August, a total of 704 cases were identified with 18 associated deaths. There are is already a limited number of health workers in the country and in particular Somali region, putting extreme strain on capacity to serve patients, especially for non-emergency care(EPHI, 2020).

Thus, COVID-19 is likely to overwhelm an already fragile health-care delivery system and reduce the availability of services for endemic health concerns such as malaria and diarrheal diseases including cholera(Poole et al., 2020). Ethiopia’s safe water supply is 30 per cent, serving as the leading cause of communicable diseases in the country.

To date, no specific antiviral treatment has been confirmed to be effective against COVID-19, and there is no vaccine so far (Cascella et al., 2020). Regarding infected patients with COVID-19, it has been recommended to apply appropriate symptomatic treatment and supportive care(Control & Prevention, 2019; Huang et al., 2020). Therefore, applying preventive measures to reduce the spread of disease is of utmost importance.

In this regard, airborne precautions and other protective measures have been discussed and proposed for prevention. Recommended preventive measures include regular hand washing with soap, social/physical distancing, and respiratory hygiene (covering mouth and nose while coughing or sneezing) (Control & Prevention, 2019; Organization, 2020c).

The WHO also issued guidelines on the use of face masks in different settings including in the community, home based care, and in the health care settings of COVID-19(Organization, 2020a). In this guideline, health care workers are recommended to use facemask such as those certified N95 when performing aerosol-generating procedures and to use medical masks when providing care to suspected or confirmed COVID-19 cases. However, these effective prevention and control practices depends on awareness and compliance among health workers at all levels.

A poor level of knowledge has been implicated in the rapid spread of the infection in health facilities (McCloskey & Heymann, 2020; Selvaraj et al., 2018) and delay of treatment(Hoffman & Silverberg, 2018), and may put patients’ lives at risk. Therefore, this study is aimed to investigate the knowledge, attitude and practice of health workers towards COVID-19 infection. The findings may be useful in recommending any remedial measures and additional interventions in the study area to improve awareness, attitude and practice among health care workers.

## Method and materials

### Study design, setting and period

An institution based cross-sectional study conducted from June to August 2020 at one public hospital and two health centers in Jigjiga town. The referral hospital is a multidisciplinary specialized teaching hospital with 351 inpatient beds and the other one is district hospital with 193 inpatient beds. Currently, it provides health services to more than one million inhabitants in its catchment area. During data collection, Karamada hospital which one of the facilities selected, was turned into covid-19 treatment center and was not accessible. Most of their staff were reassigned to other health faculties to support the provision of basic health services.

### Sample size determination

Employees that were invited to participate in this study were all health care workers employed by the public health facilities in Jigjiga city council, Somali region of Ethiopia. 686 respondents believed to have patient care or specimen contact such as physicians, nurses, midwives, health officers, laboratory professions, x-ray professionals, pharmacists/druggists, anesthetists and biomedical professionals.

### Sampling technique

All public health facilities in Jigjiga city council of Somali region were identified and all health facilities were selected. The sample size of 686 was allocated to each facility proportional to the size of health professionals who were working during data collection. The health professionals were stratified by their professions in each selected facility and all health workers were selected from each stratum.

### Inclusion and exclusion criteria

Only full-time employee (health care workers) who are potentially at high-risk (physicians, medical laboratory technologists, nurses, and midwives), available during the data collection period and who are ready to take part in the study were included.

### Data collection and quality control

A structured self-administered questionnaire was used to collect the data. The questionnaire was designed as per Ahmed M. Asaad’ study towards the Middle East Respiratory Syndrome Coronavirus (MERs CoV) and also adapted from the current interim guidance and information for healthcare workers published by the CDC, updated on March 7, 2020(CDC, 2020; Giao et al., 2020). The self-administered questionnaire consisting of socio-demographic questions, and 35 questions based on knowledge, attitude and infection control practices related to COVID-19 disease in the healthcare setting.

Information sheet and consent forms was included in the first part of the questionnaire. The study participants were requested to read the informed-consent statements before they proceeded to the next section. Study participants who unwilling to take part in the study were not forced to proceed with the questions. The questionnaires were collected daily, checked for completeness and any incomplete questionnaire the respondent were contacted for completion. In addition, timely supervision of the data collection process was done by the investigators. During collection of the questionaries precautionary measures such as wearing of masks, physical distancing was observed.

## Methods of data analysis

Data was coded and entered into Epi info version 3.5.1 software and exported into STATA version 14.1 for analysis. Knowledge and practice questions were scored as 1 or 0 for correct and incorrect responses, respectively. The total knowledge score varied between 0 (with no correct answer) and 15 (for all correct answers), and a cut off level of ≤12 was considered as poor knowledge, and >12 indicated good knowledge. The question regarding the practice was fourteen (with minimum score 14 and maximum score 42). The score of the practice based on 3 points, in which the score of 1 to 3 was given from always to never. A mean score >18.8 (answering for always) was considered as having good practice and a score of ≤18.5 indicated a poor practice (answering never or occasionally).

Whereas, attitude responses were provided with 1, 2, 3, 4 or 5 for “Strongly Agree, “Agree”, “Neutral”, “Disagree” and “Strongly Disagree”, respectively. The question regarding the attitude was six (with minimum score of 6 and maximum score of 30). The score of the attitude was based on 5 points Likert scale, in which the score of 1 to 5 was given from strongly disagree to strongly agree. A mean score >10.5 (answering for strongly agree or agree) was considered as a favorable attitude and a score of ≤10.5 was considered negative attitude (answering strongly disagree or disagree or neutral).

Summary statistics such as frequencies and proportions were computed as appropriate. Chi square test was used to compare categorical variables and ratios. T-test and ANOVA were used to analyze the relationship between the dependent (knowledge, attitude and practice), and independent variables (demographic characteristics of the participants). Spearman’s correlation was used to assess the relationship between mean knowledge and attitude scores. All the differences of estimated variables were considered statistically significant if P<0.05.

### Ethical considerations

Ethical clearance and support letters were obtained from the Ethical Review Committee (ERC) of the College of Medicine and Health Sciences, Jigjiga University. The support letter then submitted to the public health facilities. Then, permission ws obtained from the health facilities director and department/section heads. Study participants were informed about the purpose and importance of the study through written informed consent before the data collection process. In addition, participants who are unwilling to take part in the study and those who need to quit their participation at any stage were informed to do so without any restriction.

## Result

### Socio-demographic characteristics

A total of 434 heath workers completed the survey questionnaire. Of the 686 HCWs approached, total of 434 HCWs responded (response rate = 63%). A vast majority of the participants were male (n = 293, 67.5%), with a mean age of 27.6 (SD: 5.3) years and below 40 years of age (n = 423, 97.5%). Majority of the participants were working in Referral Hospital (n = 345, 79.5%). Three hundred twenty-two (74.2%) participants were nurses, 36 (8.3%) were physician, 31 (7.1%) were laboratory technician/technologist, 26(6.0%) were pharmacist, 12(2.8%) were x-ray technicians and 7 (1.6%) were dentist. Of the 434 participants, 307 (70.7%) had more than 5 years’ experience. The main sources of information about COVID-19 among participants were information from international health organizations like the CDC and WHO, Ministry of Health, Ethiopia media sites, News Media and social media such as WhatsApp and Facebook. Table 1 summarizes the sociodemographic characteristics of the participants.

**Table 1:**
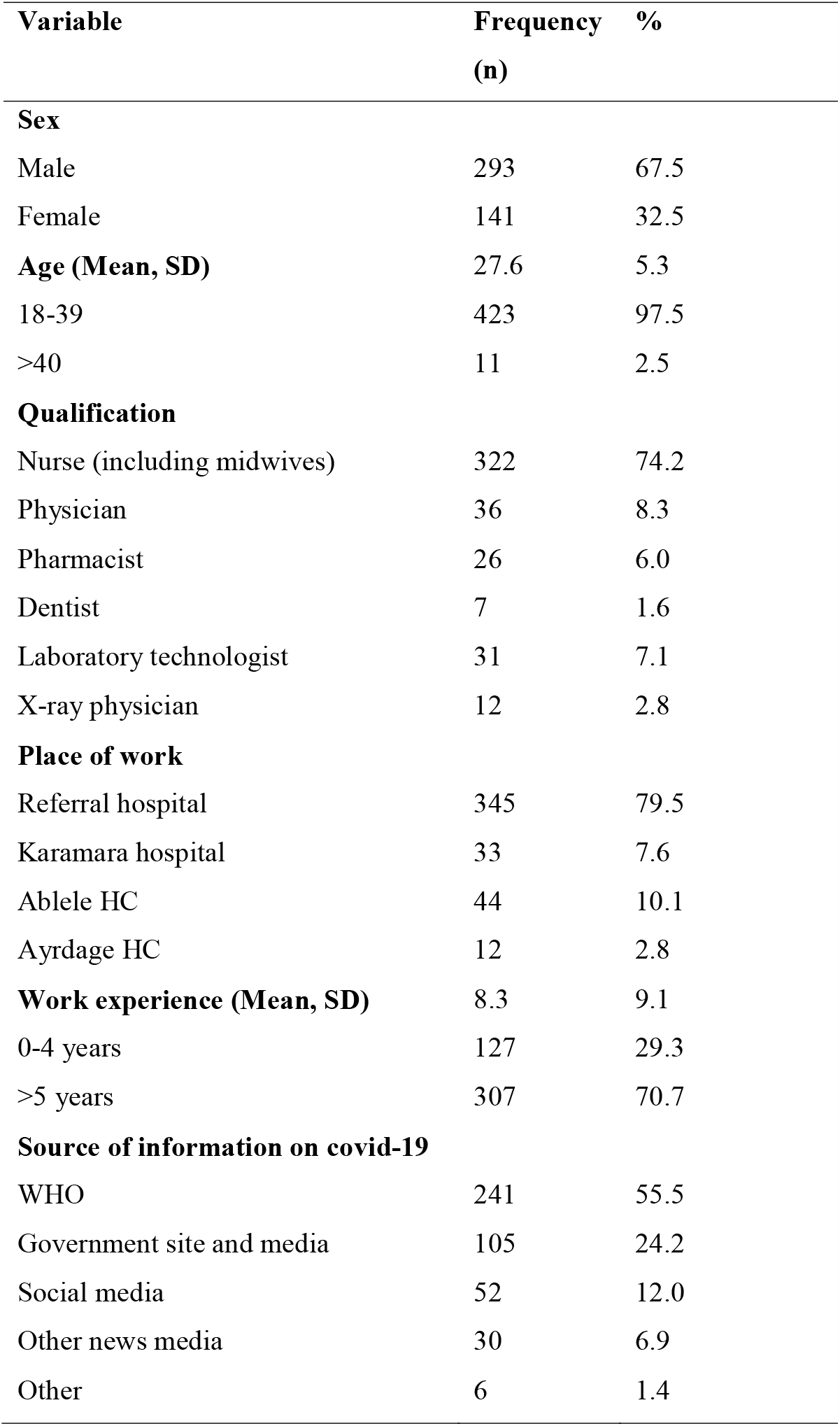
Sociodemographic characteristics of the participants.

### Knowledge

Almost all HCWs were able to correctly identify COVID-19 key symptoms. Data from current study revealed that 93% of respondents fully understood that isolation and treatment of people who are infected with the COVID-19 virus are effective ways to reduce the spread of the virus. Almost 90% were also aware of factors likely to be associated with severity of the disease.

Majority of the respondents understood dynamics of COVID-19 infectiousness: 91.5% respondents were aware of possibilities to infect before the onset of symptoms; 90.8% of HCWs responded true on droplets as a major transmission route. Considerable proportion of respondents (93.1%) were informed that the incubation period is not constant and could vary from 2-14 days.

### Attitude

The mean attitude score 10.5 (SD: 4.1). Overall, there was poor attitude among HCWs toward COVID-19. Only 45.2 % (n = 196) of the participants had a good attitude toward COVID-19. Only 48.8% (n = 212) were confident that COVID-19 can be controlled by public health institute (Table 3). When asked about the using mask, up to 49.3% (n = 214) believed that It is imperative to use surgical mask when working with patient with COVID-19.

**Table 2:**
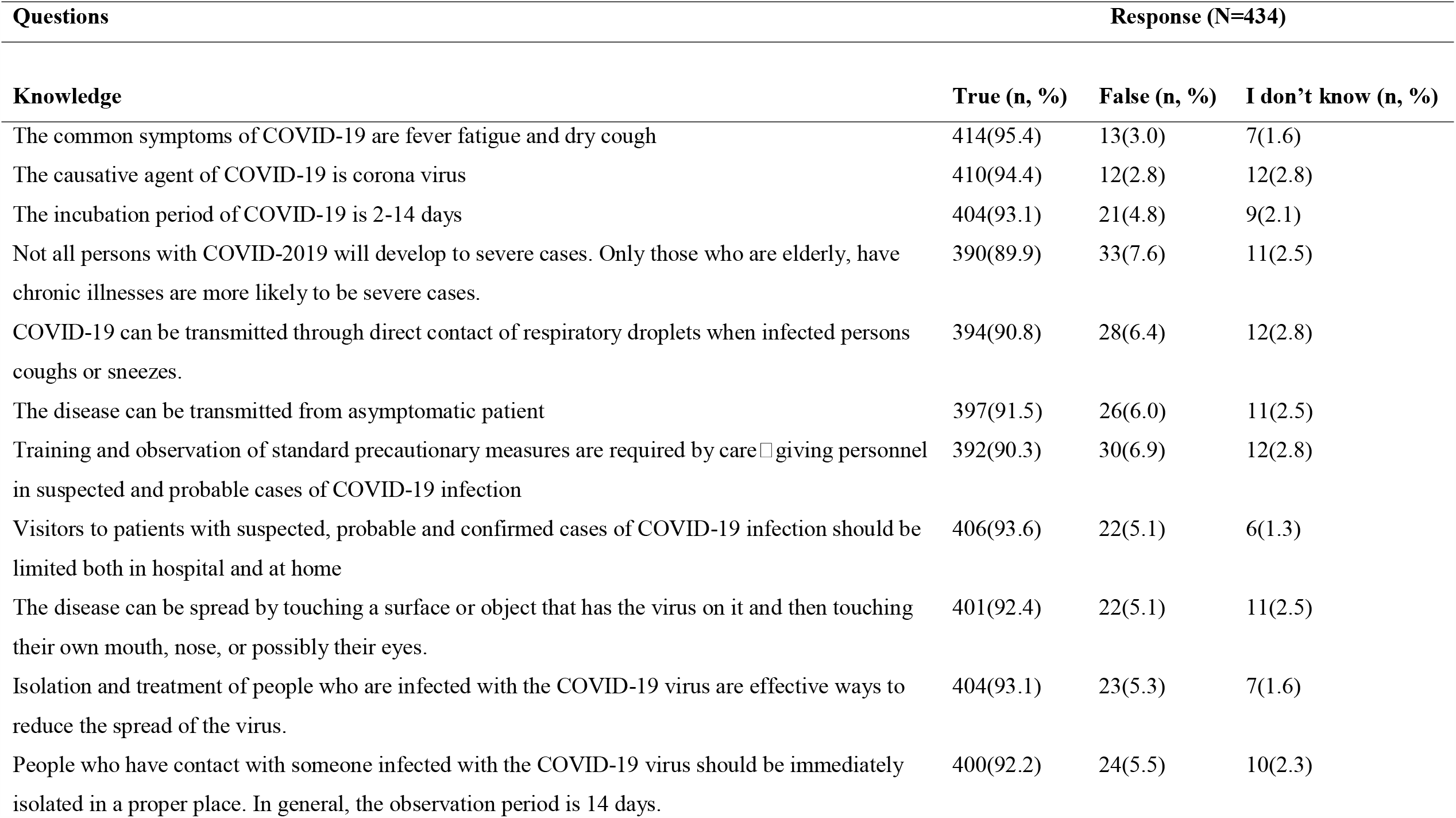
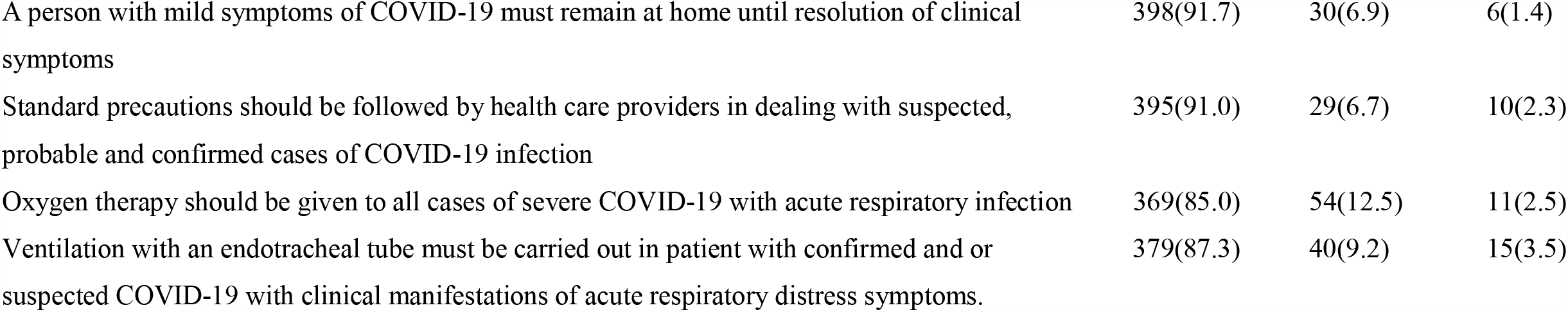
Knowledge of HCWs toward the COVID-19, 2020.

**Table 3:** Attitude of HCWs toward the COVID-19, 2020.

| Questions | Response (N=434) |  |  |  |  |
| --- | --- | --- | --- | --- | --- |
| Attitude | SA | A | N | D | SD |
| Caring for patients with COVID-19 infection may be a threat to health care personnel | 226(52.1) | 147(33.9) | 28(6.4) | 24(5.5) | 9(2.1) |
| Public health agencies like EPHI can control Outbreak of COVID-19 | 212(48.8) | 121(27.9) | 42(9.7) | 50(11.5) | 9(2.1) |
| COVID-19 can have a negative effect on the economies of the countries involved | 231(53.2) | 135(31.1) | 24(5.5) | 36(8.3) | 8(1.9) |
| It is important to report suspected cases to health authorities | 301(69.3) | 104(24.0) | 11(2.5) | 13(3.0) | 5(1.2) |
| COVID-19 is preventable | 226(52.1) | 136(31.3) | 33(7.6) | 23(5.3) | 16(3.7) |
| It is imperative to use surgical mask when working with patient with COVID-19 | 214(49.3) | 136(31.3) | 23(5.3) | 26(6.0) | 35(8.1) |

### Practice

About practical measures put in place by HCWs in order to protect themselves and their families, 72% of health care workers reported to always wore personal protective equipment when coming into contact with the patients and up to 67.5% washed their hands before and after touching each patient. Other measures included: avoiding going to crowded places and avoiding shaking hands, hugging or kissing. Unfortunately, as high as 72.6% (n = 315) of the participants had avoided patients with symptoms suggestive to those of COVID-19 (Table 4).

**Table 4:**
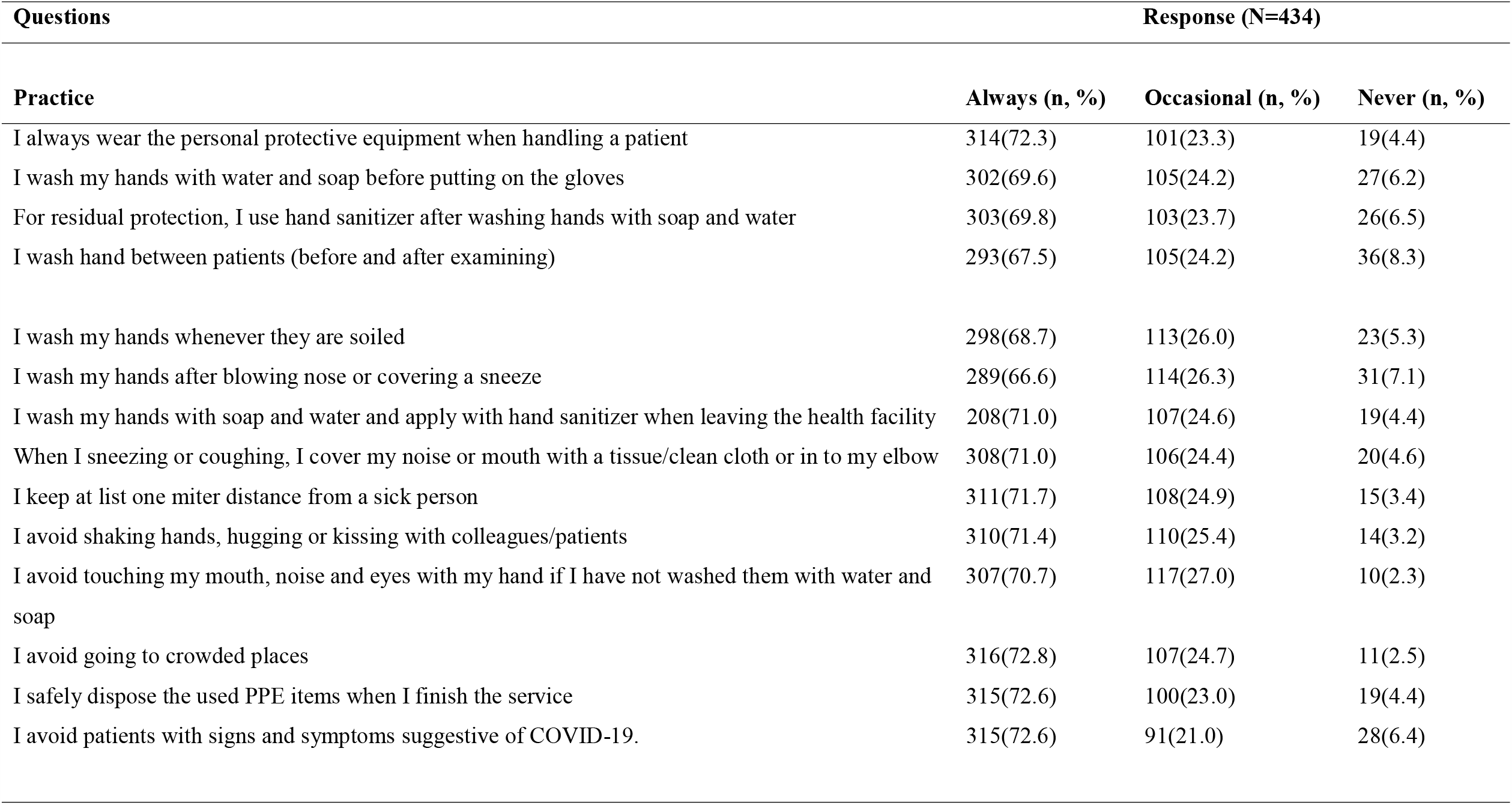
Practice of HCWs toward the COVID-19, 2020.

### Comparison of KAP between demographic characteristics of HCWs

Almost ninety percent (n = 381) of the participants scored 12 or more and were considered to have su□cient knowledge. Of the total participants, more male participants had sufficient knowledge than those of female participants (89.8 vs. 83.7%), however this di□erence was not statistically significant (p = 0.07). More participants working at the referral hospital had sufficient knowledge than other health facilities (p<0.001).

Only 45% of the participants had positive attitude (n=196). Bivariate analysis revealed that more HCWs working at Ayardage HC had good attitude compared to those HCWs working at other health facilities (p=0.02). More HCWs who used government media like television to access information on COVID-19 had better attitude (p = 0.009). There was no statistically significant difference between attitude and other sociodemographic variables (Table 5).

**Table 5:**
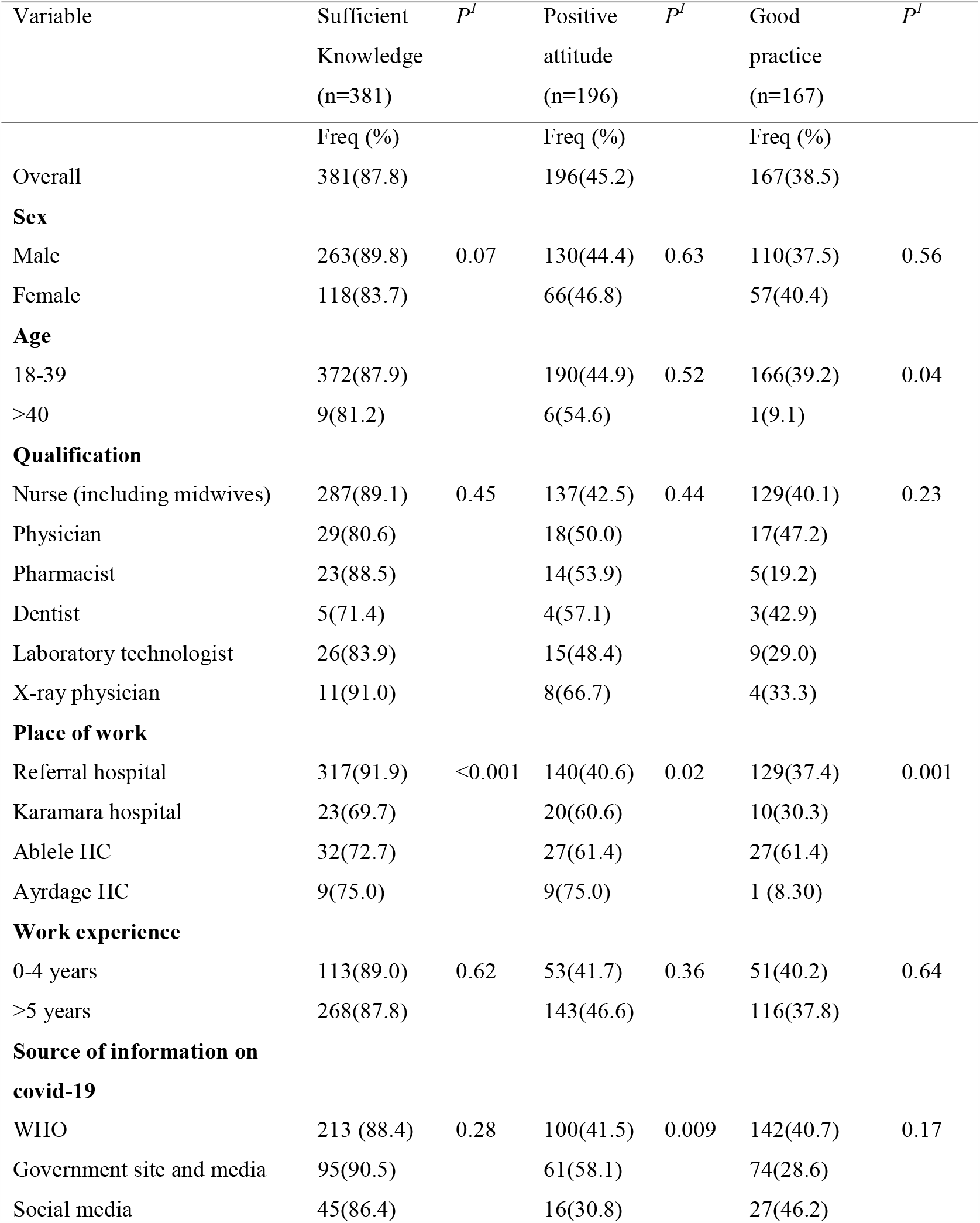
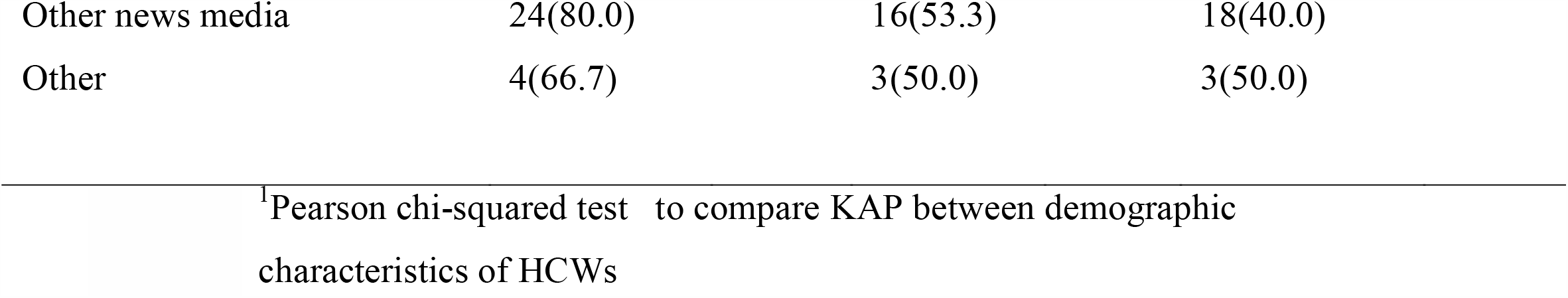
Comparison of COVID-19 knowledge, attitude and practice between different groups.

Overall, up to 38.5% (n = 167) of the participants had good practices (mean score ≥ 18.8). HCWs ≤40 (p=0.04) and those working at Ayardage HC practiced more than their counter parts (p <0.001) (Table 5).

### Distribution of KAP

The mean knowledge, attitude and practice score was 13.7 (SD: 2.6), 10.5 (SD:4.1) and 18.8 (SD:5.8) respectively. Male participants had higher knowledge score than female (p=0.013). Similarly, nurses had higher knowledge score compared to other health professionals (p=0.04). The level of knowledge among the healthcare workers working at Referral hospital (p<0.001) and those who seek covid-19 related information from world health organization (p<0.001). (Table 6).

**Table 6:**
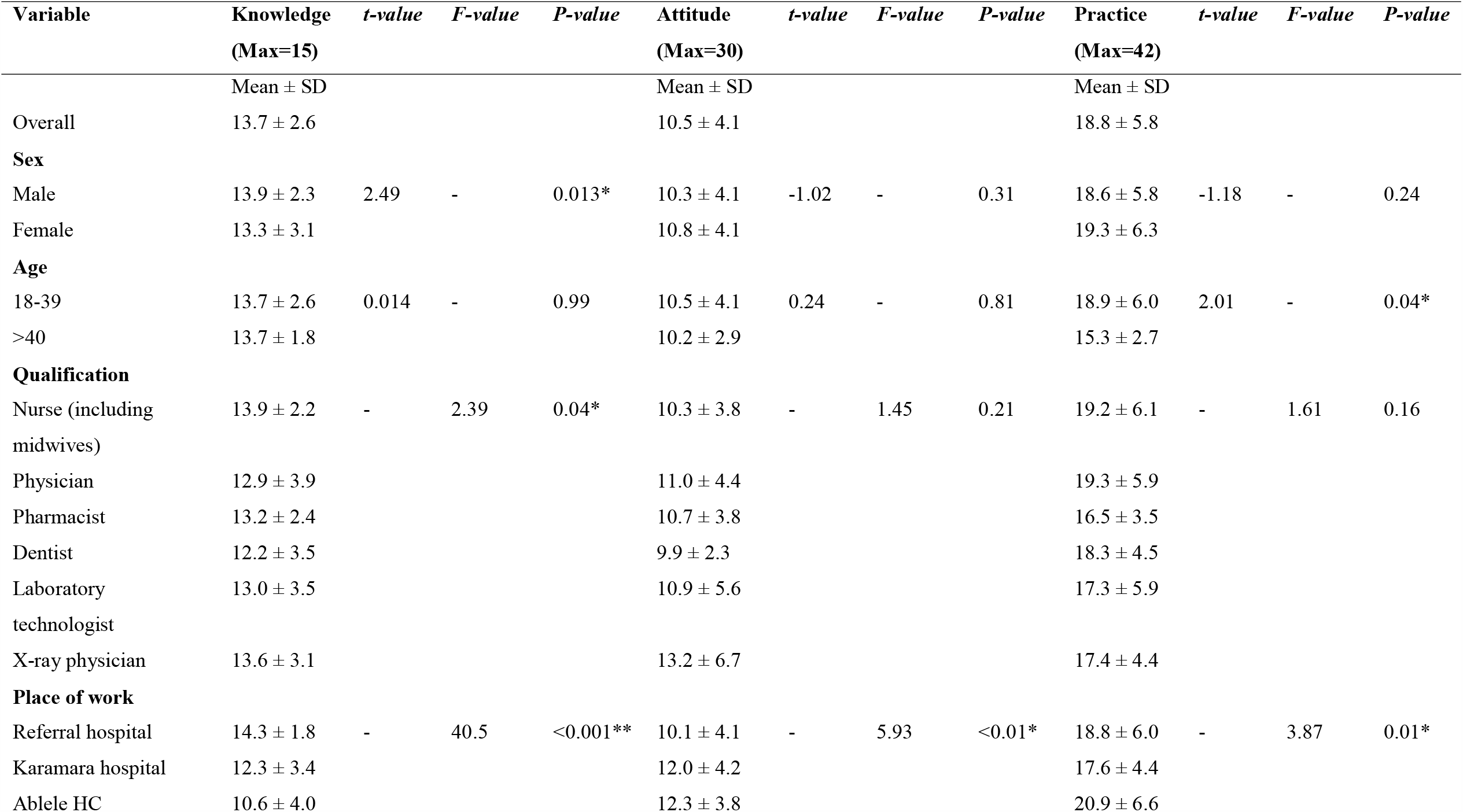
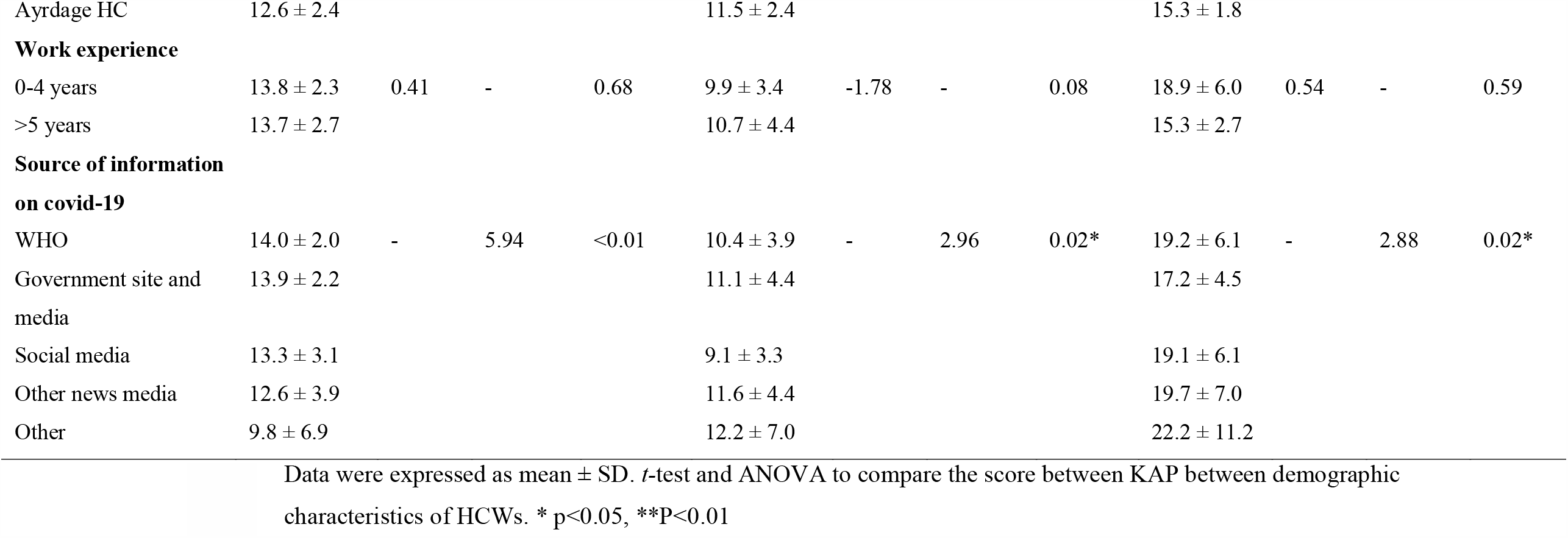
Distribution of knowledge and attitude scores among healthcare workers, 2020.

**Table 6:**
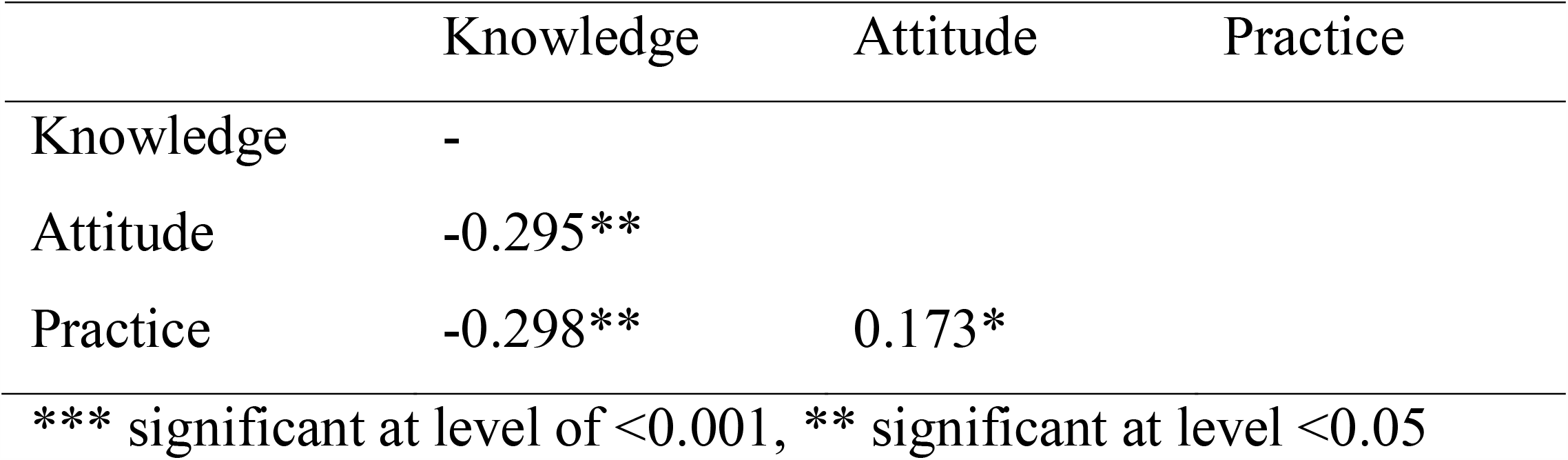
Correlation between knowledge, attituded and practice.

|  | Knowledge | Attitude | Practice |
| --- | --- | --- | --- |
| Knowledge | - |  |  |
| Attitude | -0.295** |  |  |
| Practice | -0.298** | 0.173* |  |

\*\*\* significant at level of $< 0.001$ , \*\* significant at level $< 0.05$

Attitude score was higher among health workers working at Ablele HC and among HCWs who seek information from other sources. Likewise, practice score was higher among younger professional (p=0.04), those working at Ablele hospital (p=0.01) and those who seek covid-19 information from other sources (p=0.02). (Table 6

### Correlation between knowledge, attitude and practice

Table 5 presents the correlation coefficient between whole grains knowledge, attitude and practice. It was found that attitude and practice domain were inversely associated knowledge score (r=-0.295, p <0.01) and (r=-0.298, p <0.01) for attituded and practice respectively. Significant positive correlations were found between attitude with attitude (r=0.173, p<0.0003).

## Discussion

The current work assessed the knowledge, attitude and practice toward the COVID–19 outbreaks among HCWs working at public health facilities. In this study, the socio-demographic, knowledge level, attitude, and infection prevention practical level response of 434 HCWs were analyzed.

Our study found that most of the HCWs were well informed with COVID-19 related knowledge, during the outbreak. Among these HCWs, the level of knowledge about COVID-19 was higher male participants and nurse. From our study, 87.8% of HCW had su□cient knowledge about COVID-19 which is higher than values reported by studies conducted in Uganda and Northern Ethiopia(Olum et al., 2020; Tadesse et al., 2020) where 69% and 74% of HCWs had sufficient knowledge. The proportion of HCW with sufficient knowledge in the current study is similar to those studies conducted in China(Huynh et al., 2020; Zhou et al., 2020).

In our study, most of the participants used information from international and governmental media (websites and verified social media pages). Our study suggests that majority of HCWs with sufficient knowledge on COVID-19 used news media such television. This suggests that such media should be frequently used to disseminate information on COVID-19 by the stakeholders involved in covid-19 response.

About 52% of HCW believed that caring for COVID-19 patients may be a threat to heath care workers which is similar to findings by study conducted in China(Zhou et al., 2020) which showed around 85% of the surveyed HCWs were afraid of getting infected while caring for their patients. HCWs help patients during their routine tasks such as patient consultation, infusion, dressing changes and surgery. They must also deal with many other emergency situations, and they may become infected with the virus if they are not cautious.

About 14% of HCW believed that wearing general medical masks may not protect the spread COVID-19 contrary to findings by Ng et al. which showed adequate protection(Ng et al., 2020).

Our study reveals that majority of HCWs have a negative attitude toward COVID-19 which is in congruence with a KAPs study on COVID in Uganda among HCWs(Olum et al., 2020) but in contrast to study in China on COVID-19 and Ethiopia (Jemal et al., 2020; Zhou et al., 2020). However, Spearman’s analysis found that a significant negative correlation between the mean knowledge and attitude scores of HCWs about COVID-19 (r=-0.295, P<0.001) which is in consistent with a study conducted in Vietnam(Huynh et al., 2020). In other words, the lower the attitude scores were, the higher the probability of positive attitudes; while the higher the knowledge scores were, the higher the probability of good knowledge. Therefore, a good knowledge COVID-19 was directly associated with a positive attitude.

Our study shows that majority of HCWs have low score of COVID-19 prevention practices contrary to findings by other studies on coronaviruses(Olum et al., 2020; Tadesse et al., 2020; Zhou et al., 2020). Though majority of the HCWs are following key infection prevention and control practices recommended by the Ministry of Health Ethiopia and WHO. These include regular hand hygiene, social distancing and wearing personal protective equipment when in high risk situations. Seventy two percent and 67.5% of HCWs reported wearing personal protective equipment when in contact with patients and washing hands before/after handling patients. These are very vital practices to prevent transfer of COVID-19 from patients to patients and to the HCWs themselves. However, up to 73% of HCWs admitted having avoided patients with symptoms suggestive of COVID-19. This can be attributed to shortage of personal protective equipment globally (Bauchner et al., 2020; Ranney et al., 2020).

Our study has some limitations. Firstly, no standardized tool for assessing KAPs on COVID-19 has been previously validated. We have however adapted and modified a previously published tool for assessment of KAP toward prevention of respiratory tract infections. Secondly, only HCWs in public health facilities in parts of Somali region were surveyed and the results of this study may not reflect the KAPs of HCWs in the private sectors. A similar study may be extended to the community.

In conclusion, we found that more than 85% of HCWs have su□cient knowledge on the transmission, diagnosis and prevention of the transmission of COVID-19. Knowledge on COVID-19 was significantly higher among HCWs who are working at referral hospital. There was statistically significant di□erence in the level of knowledge about COVID19 among health care workers. About 55% of the respondents had poor attitude toward COVID-19 and just over 38% of the HCWs had good practices toward COVID-19 especially those younger than 40 years. In nutshell, the overall level of knowledge was good. However, the attitude and practice were relatively low. We therefore, recommend strategies for enhancing the capacity of healthcare workers to develop positive attitude and practice.

## Data Availability

The datasets for this study is available from the corresponding author on reasonable request.

## List of abbreviation

CDC: Centre of Disease Control
CI: Confidence Interval
EOC: Emergency Operating Centre
EPHI: Ethiopia Public Health Institute
ERC: Ethical Review Committee
HCW: Health Care Worker
IPC: Infection Prevention Control
PPE: Personal Protective Equipment
WHO: World Health Organization

## Acknowledgment

We would like to extend a special thanks to Jigjiga University for funding researches on covid-19 which may be used in planning health programs about the emerging viral disease.

## Declarations

### Availability of data and materials

The dataset for this study is available from the corresponding author on reasonable request.

### Ethical approval

Ethical clearance and support letters were obtained from the Ethical Review Committee (ERC) of the College of Medicine and Health Sciences, Jigjiga University. The support letter then submitted to the five public health facilities. Then, permission obtained from the health facilities director and department/section heads. Study participants were informed about the purpose and importance of the study through written informed consent before the data collection process. In addition, participants who are unwilling to take part in the study and those who need to quit their participation at any stage were informed to do so without any restriction.

### Consent for publication

Not applicable

### Authorship

AM conceived the study, prepared the proposal, analyzed the data, interpreted the findings and wrote the manuscript. TY, MO, MA, OM, MA and AB were involved in developing the study proposal, data analysis and reviewing the manuscript.

